# *“We are their eyes and ears here on the ground, yet they do not appreciate us”* - Factors influencing the performance of Kenyan community health volunteers working in urban informal settlements

**DOI:** 10.1101/2023.03.22.23287562

**Authors:** Michael O Ogutu, Eric Kamui, Timothy Abuya, Kui Muraya

## Abstract

Community Health Volunteers (CHVs) play a crucial role in linking the community with the formal health system, particularly in low- and middle-income countries. Studies in Kenya have focused on the implementation of the Kenya Community Health Strategy (CHS) in rural, nomadic, and peri-urban areas; with limited information on the factors that influence CHV performance in urban informal settlements. This study therefore explored factors that influence CHV performance in urban informal settlements within Nairobi Kenya and ways in which CHVs can be supported to enhance their wellbeing and strengthen community strategies. The study was undertaken in two urban informal settlements within Nairobi County. Thirteen focus group discussions (total of 123 participants) and three key informant interviews were conducted with a range of respondents. Various topics covering the design of the Community Health Strategy (CHS) and broader contextual factors that affect CHVs’ performance, were discussed and the data analysed using a framework analysis approach. The key programme design factors identified as influencing the performance of CHVs working in urban informal setting included: CHV recruitment; training; the availability of supplies and resources; and the remuneration of CHVs. Health system factors that influenced CHVs performance included: nature of relationship between healthcare workers at local referral facilities and community members; the availability of services and perceived corruption at the referral facilities; and CHV referral outside of the local health facility. Whereas the broader contextual factors that affected CHV performance at the community level included: demand for material or financial support; perceived corruption in community programmes; and neighbourhood insecurity. These findings suggest that like other CHVs working in both the rural and peri-urban settings, CHVs working in urban informal settlements in Kenya face a myriad of challenges that impact on their wellbeing and work performance. Therefore, to enhance CHVs’ well-being and improve their performance, the following should be considered: adequate and timely remuneration for CHVs, appropriate holistic training, adequate supportive supervision, and ensuring a satisfactory supply of resources and supplies. Additionally, at the health facility level, healthcare workers should be trained on appropriate and respectful relations with both the community and the CHVs, clarity of roles and scope of work, ensure availability of services, and safeguard against corrupt practices in public health facilities. Lastly, there is a need for improved and adequate security measures at the community level, to ensure safety of CHVs as they undertake their roles.

## Introduction

Community health workers (CHW) are an important part of the health system, particularly in low- and middle-income countries (LMICs); and play a crucial role in linking the community with the formal health system. Different terms are used to refer to CHWs depending on context (1,2). For example, in India, they are referred to as Accredited Social Health Activists (ASHAs) while in other contexts they are known as lay health workers, traditional birth attendants, lay counsellors. For this paper, we broadly refer to them as community health volunteers (CHVs), which also aligns with the terminology used in Kenya.

Literature from LMICs shows that when involved efficiently, CHVs can improve access to primary health care services including: maternal and neonatal health services (3,4); psychosocial support services such as counselling, to individuals experiencing mental health problems like anxiety and depression (5,6); and the provision of preventive health education on topics such as hygiene and sanitation (7), exclusive breastfeeding (8,9) and family planning (10). CHV programmes also promote the provision of services such as child immunization (11), and in responding to major health outbreaks as was observed during the Ebola outbreak in West and Central Africa (12); and more recently with the COVID-19 pandemic in Kenya (13). Thus, CHVs are important actors in ultimately improving health outcomes, particularly in low-resource settings (14–18).

A systematic review by Kok et al., showed that the success of CHV programmes depends on three key aspects: programme design factors, health system factors, and broad contextual factors surrounding the workings of such programmes as shown in figure 1(2,19,20). This work was guided by the Kok et al., (2015) framework (figure 1). According to Kok and colleagues, programme design factors are features of the programme or intervention design that have a direct influence on CHVs’ performance and can be directly modified and adjusted to motivate maximum CHV performance (20). They include renumeration, recruitment, supervision and workload, community and health system links, and resources and logistics (2). Contextual factors relate to the setting in which the CHVs are working. That is, the broader environment, community, and economy; and the health system policy and practice (21). Health system factors on the other hand include issues related to human resources for health policies, service delivery, financing, and governance (20).

**Figure 1:**
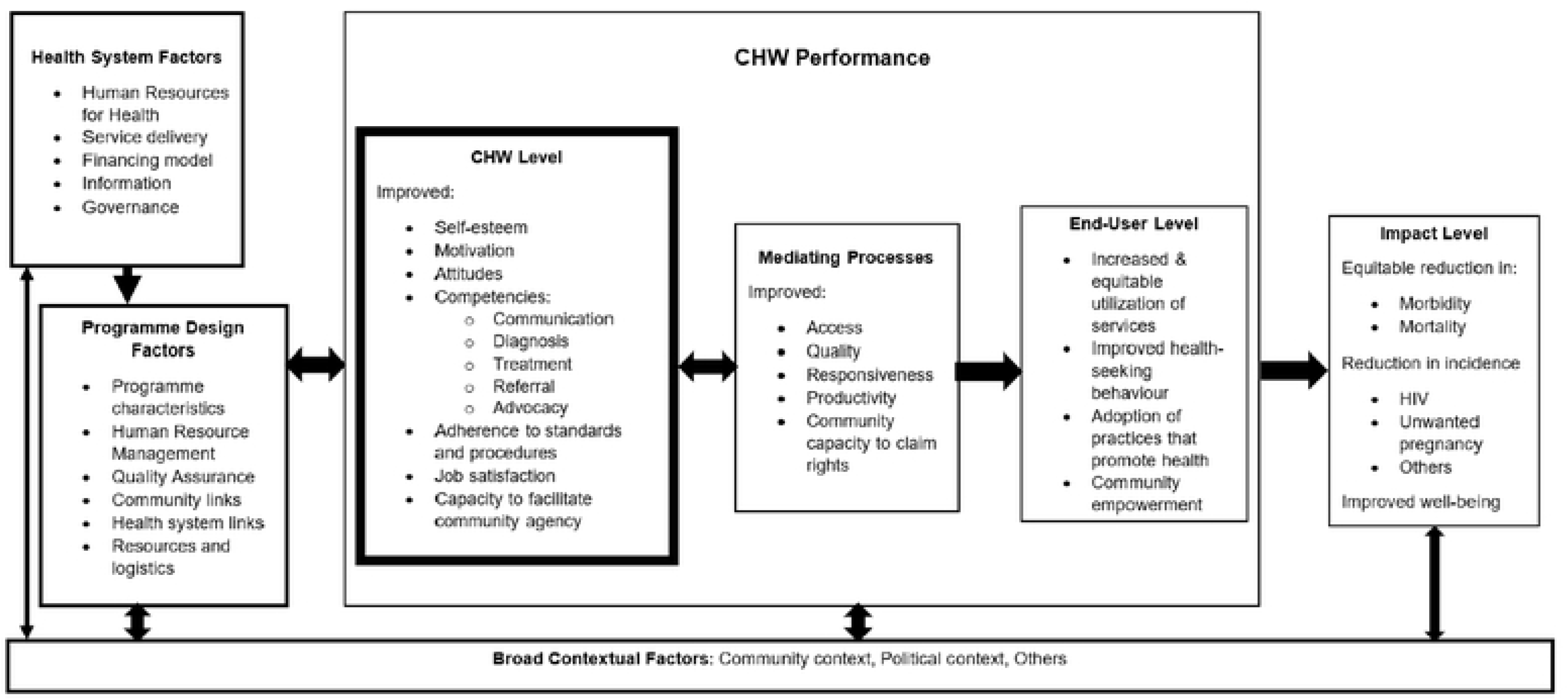
Kok et. al (2015) framework.

CHV performance can be measured at two levels, the individual (CHV) level and at the end-user (community) level (20). Attributes that are measured at the individual level include the cognitive, affective, and behavioural changes in the CHV such as self-esteem, attitude, motivation, job satisfaction, competencies/knowledge levels and guideline adherence. Whereas at the end-user or community level, one can measure CHV- attributable outcomes in the community. Such outcomes include for instance; credibility or trust in the CHVs, increased use or awareness of a health service, or the acceptance and use of services provided at a health facility following referral by a CHV (20). This work focused on assessment at the *individual* level as highlighted in bold in figure 1.

Literature shows that context matters when it comes to the performance of CHVs. It is, therefore, important to understand factors influencing CHV performance in specific contexts such as urban informal settlements (UIS). For example, in addition to the factors highlighted above, a systematic review by Ogutu et al., (22) showed that specific to UIS in LMICs, some of the factors that negatively impacted on the performance of CHVs at the broader contextual level in UIS included insecurity and the demand for material or financial support by households.

In Kenya, it is estimated that more than half of the urban population lives in informal settlements, with 56% residing in the capital city Nairobi (23). These populations are often characterized by a high burden of disease and limited access to health care (24–28). To increase health care access in these and other settings, in 2006 the government rolled out the Kenya Community Health Strategy (CHS). In brief, this strategy predominantly relies on CHVs to deliver a range of services, and act as a link between the formal healthcare system and the community (30–33). The CHVs act as a link to the primary health facilities through government-employed Community Health Assistants (CHAs), formerly known as Community Health Extension Workers (CHEWs). CHAs are trained health workers linked to a specific primary health care facility and provide support and supervision to an assigned number of CHVs.

Current Kenyan studies on CHVs have focused on nomadic, rural, and peri-urban areas with less attention given to UIS (34–41). Drawing on the Kenya CHS as the ‘intervention’ or ‘programme’ under implementation; this study therefore aimed to explore factors that affect CHV performance in UIS within Nairobi, Kenya. This study also aimed to identify and explore ways in which CHVs can be better supported to enhance their wellbeing and strengthen community strategies.

## Methods

### Study sites

This qualitative exploratory study was conducted in two UIS (Kibera and Mathare) of Nairobi County. Kibera also known as Kibra is one of the largest informal settlements in East Africa (42), and is one of the 17 sub-counties within Nairobi County. It is located approximately 5 km southwest of the Nairobi central business district (CBD) and occupies 12.1km^2^. It is sub-divided into 14 sub-sections commonly referred to as ‘villages’ as indicated in figure 2 (43,44). Kibra sub-county is mainly inhabited by the Luo, Luhya, Kisii, Kamba and Kikuyu ethnic communities (25,42). There are two major hospitals located within its environs (Mbagathi Sub-County Referral Hospital and Kenyatta National Hospital). The area is also served by several other smaller government, private and non-government organizations (NGO) health centres, dispensaries, clinics, and nursing homes.

**Fig. 2.**
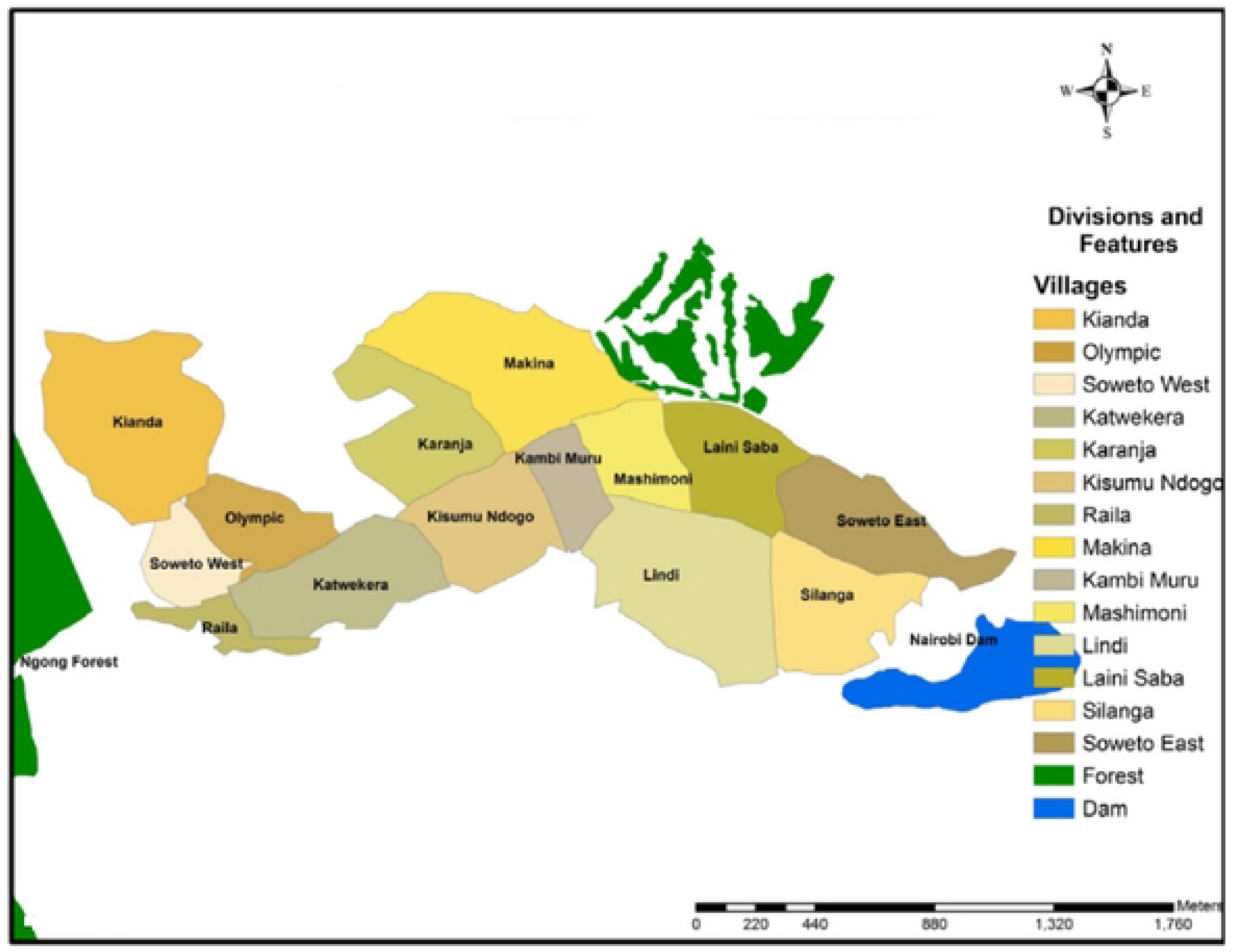
Map of Kibera urban informal settlement, adapted from Mutisya et al., 2011(43)

Mathare is the second largest informal settlement in Nairobi after Kibera (45) and is also a sub-county. It is located northeast of the Nairobi CBD and has 16 villages in total as indicated in figure 3. Mathare sub-county is mainly inhabited by the Luo, Luhya, Kikuyu, Kisii, and Kamba ethnic communities (42). Like Kibera, Mathare is also located near a major government health facility (Mathare Mental National Referral Hospital). The area is also served by several smaller government, private and national and international NGO health facilities.

**Fig. 3.**
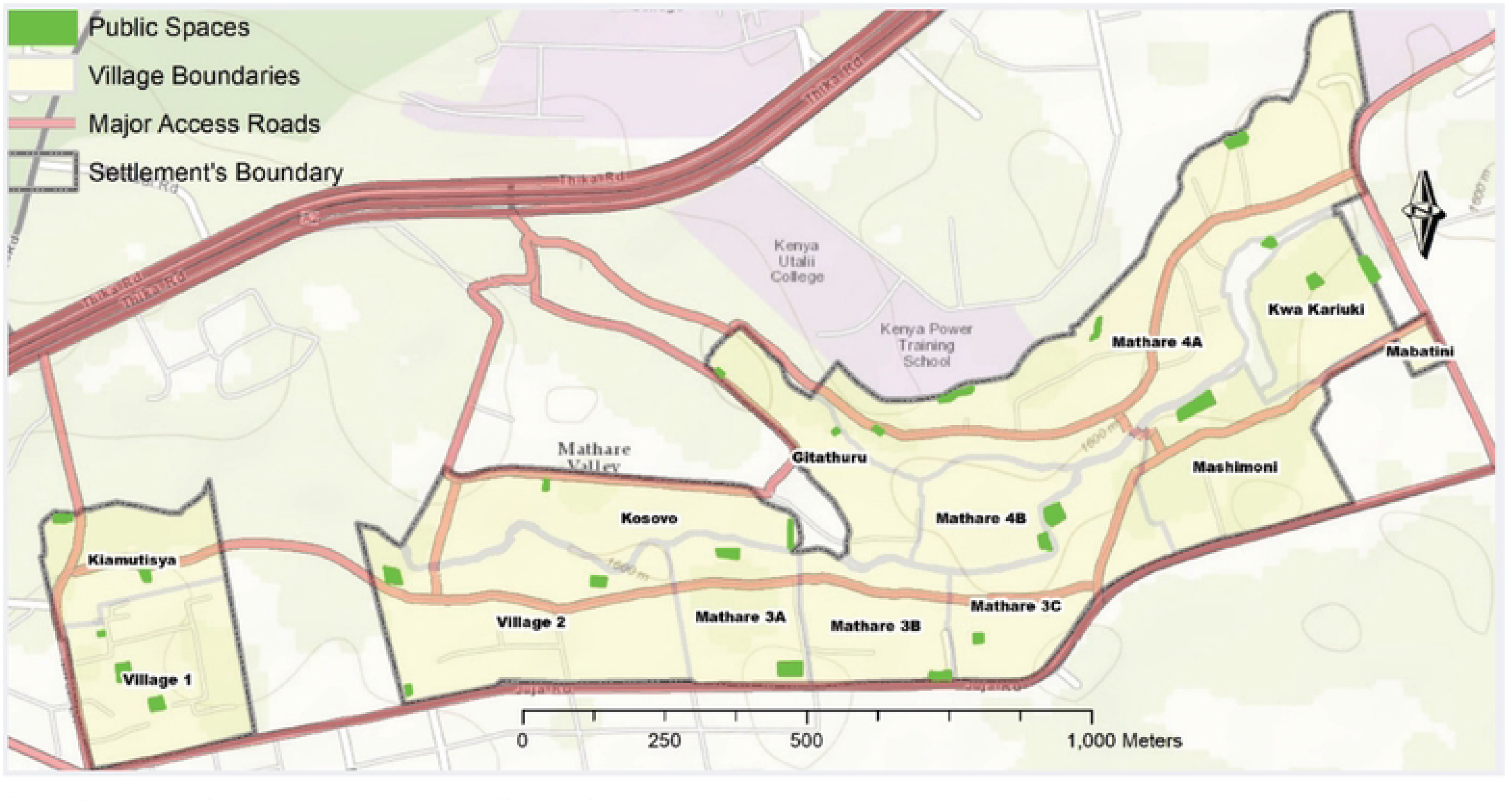
Map of Mathare urban informal settlement, adapted from UN-Habitat., 2020(45)

### Data collection and analysis

Data was collected using key informant interviews (KIIs) and focus group discussions (FGDs). KIIs (involving two sub-county coordinators and one CHA) were conducted in English, whereas the FGDs (involving the rest of the participants) were conducted in both Kiswahili and English. All discussions were audio-recorded and thereafter transcribed verbatim and translated where needed. The lead author verified all the translated transcripts. During the transcription and translation process, the lead author also counterchecked several randomly selected transcripts against the raw audio recordings; to check for accuracy and quality.

Following every FGD, the first and second author who collected the data had a debrief session where they developed detailed summary sheets, and discussed any pertinent issues or dilemmas emerging from the field; including those that needed to be taken into account for subsequent FGDs. The last author also joined most of these debrief sessions. In addition, the lead author developed detailed summary sheets after every KII. These summary sheets were used to help manage the data, as well as to keep whole the ‘story’ from each FGD or KII, prior to the process of splitting the data into themes (coding). Following this, the data were analysed using a framework analysis approach (46). This involved: extensive familiarization with the data (‘immersion’ in the data) by reading and re-reading of transcripts, listening to audio-recordings, and re-reading the developed summary sheets; consultatively developing a coding framework based on the study objectives and preliminary emergent themes; and coding the entire dataset using NVivo software, to search for further emergent themes. Comparison charts were thereafter developed to identify patterns and explore relationships between the different factors that influenced the performance of CHVs. The lead author also kept a reflective journal where he documented events both during and after field visits. In this journal the author reflected on how his own beliefs and assumptions might have influenced data collection, analysis and interpretation; and noted down emerging dilemmas and potential solutions.

Ethical approval to conduct this study was obtained from the KEMRI Scientific and Ethical Review Unit (KEMRI/SERU/CGMR-C/228/4190). Written informed consent was obtained from all participants prior to data collection. The different dates for recruiting the different study participants are indicated in the table below.

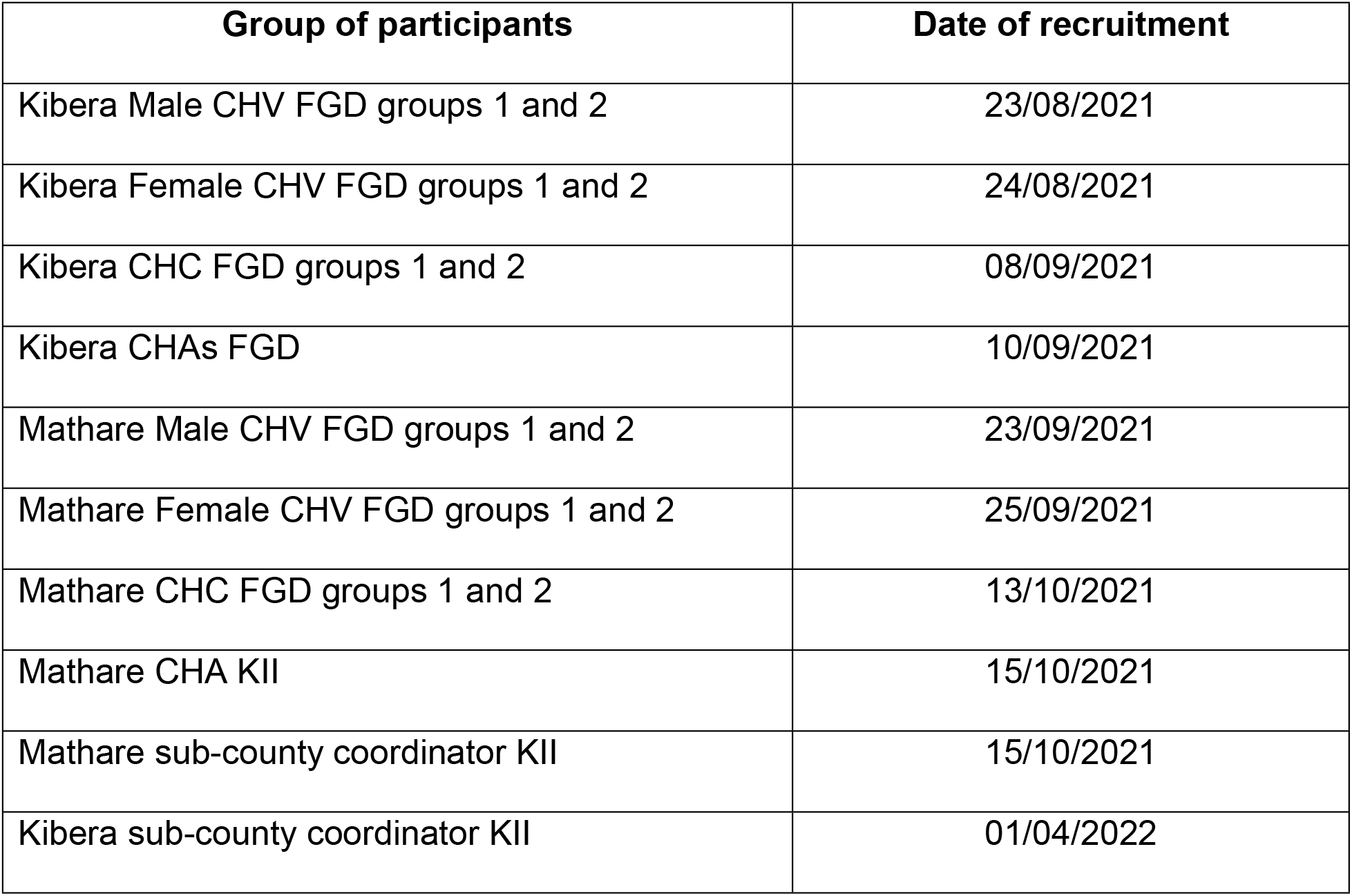

## Results

This section begins by presenting the general characteristics of the study participants, followed by the factors that influenced the CHVs’ performance as outlined in the Kok*.et al.,* (2015) framework.

### General characteristics of study participants

The study involved 126 participants, including two sub-county officials, 14 community health assistants, 77 CHVs and 33 community health committee (CHC) representatives. The participants varied in age ranging from 22 to 63 years old. Of the 77 CHVs, 16 had primary school education, 34 secondary school education, and 27 had tertiary level education (24 diploma and three university level education). Length of service varied from one year to as many as 25 years in service. Table 1 below gives additional information on the study participants’ characteristics.

**Table 1:** Characteristics of study participants.

| Category of participants | No. of participants | Age - range | Level of education | Length of time in service |
| --- | --- | --- | --- | --- |
| CHS sub-county coordinators | 2 | 24 - 49 | - | 2 – 3 years |
| CHAs | 14 |  | - | 7 months – 11 years |
| CHVs | 77 |  |  | 2 – 16 years |
| Male | 38 | 22 - 62 | Primary (6) |  |
|  |  |  | Secondary (12) |  |
|  |  |  | Diploma (19) |  |
|  |  |  | University (1) |  |
| Female | 39 | 24 - 57 | Primary (10) | 1 – 25 years |
|  |  |  | Secondary (22) |  |
|  |  |  | Diploma (5) |  |
|  |  |  | University (2) |  |
| CHC representatives | 33 | 30 - 63 | - | 1 – 20 years |

### Programme design factors

As earlier described, for this study, the Kenya CHS was the programme or intervention under consideration, and the factors described below are related to its implementation. In this context, the programme design factors that were observed as being important and impacting on CHV performance included CHV recruitment/selection, training, supportive supervision, supplies & resources, and CHV remuneration. Each of these are discussed in turn below.

#### CHV recruitment

CHV recruitment was done through Community Health Committees (CHC), especially when forming new Community Health Units (CHU). Both CHCs and CHUs are described elsewhere – see details under supplementary information. The CHV positions were advertised in the community by word of mouth and through public meetings (*barazas*) and those interested applied through the local administration office. The area chiefs who are also members of the CHC, then called for a *baraza* where the applicants were vetted. According to the respondents and as illustrated below, for one to qualify to be a CHV, they had to be at least 18 years of age, be a good role model in the community, be willing to volunteer, be a resident of the local area, and know how to read and write as illustrated below.

> *“…CHVs are recruited in a chief’s baraza… the first step during recruitment is normally to come up with the Community Health Committee… and thereafter the CHV positions are advertised… and any person interested in the position is asked to come to the baraza. Once the baraza is called, the Community Health Committee members are then given the qualifications… like one, the CHVs must be 18 years and above…be role models…be able to read and write because there are reports that are going to be required. The other thing is that they must be willing to volunteer their services…the community members are also involved in the vetting process because we’ll ask them whether they approve of them [the CHVs] or not. So, it is done in such a way that all villages are represented…”*
>
> — (sub-county official)

The voluntary nature of the CHV positions had a negative impact on the recruitment exercise since only those who had freshly graduated from secondary school applied; and thereafter dropped out of the CHV role once they secured paid opportunities. In other words, many viewed the role as a stopgap measure while they searched for paid employment. Consequently, the CHC members recommended the CHV positions to community members who were personally known to them, as opposed to publicly advertising the positions.

#### CHV training

All CHVs were supposed to receive two types of training, basic and technical training. According to the participants, the basic training focused on the Kenya CHS and highlighted the roles and responsibilities of the CHVs. Whereas the technical training comprised of technical modules such as integrated community case management, and water and sanitation. Majority of the CHVs reported receiving only basic CHV training upon being recruited. This was because the technical training was offered by various non-governmental partners and was dependent on the project goals of implementing partners. This also meant that the CHVs who received technical training were those situated in the localities where programs were being implemented by the non-governmental partners (in turn disadvantaging those from other areas). Other factors that were considered in selecting CHVs for technical training included; number of slots available for a particular training, CHV’s level of activeness, ensuring there was rotation in who was selected for training , ability of a CHV, and personal preference by Community Health Unit leaders. Furthermore, since the CHVs were not remunerated for their work and to help keep them accountable for their performance, the trainings were sometimes used to ‘reward’ those CHVs considered to be active in the community as explained below by a CHU leader.

> *“…I do select [people for training]. I am the chairman of a CHU…mostly those who are picked for trainings are those seen to be active. Since there are others who if you ask them to bring their reports … they will give you like 10 excuses [why they can’t]. So, I cannot call a person like that for a training or even an activity…”*
>
> — (Male CHV, unit leader).

Most of the CHVs felt that the trainings empowered them with basic knowledge and skills to perform their roles confidently. The technical training also brought change of attitude among the CHVs, with for example, some male CHVs becoming more empathetic as highlighted below.

> *“I [previously] didn’t care about the community. But after I got the training, it changed my attitude and I started to appreciate other people…I now feel for others and whenever I meet a sick person, I try to assist and even give a referral in case one is needed.”*
>
> — (Male CHV, Kibra)

Despite their positive impact, the trainings were sometimes used to ‘punish’ those CHVs who were viewed as inciters, for example those who asked for better working conditions. This in turn resulted in affected CHVs becoming demotivated and preferring to engage in private income generating activities to provide for their families. Selection for technical training was also linked to potential favoritism, which led to CHV demotivation as illustrated in the quote below.

> *“…Sometimes when trainings come, some people will go for more trainings and others are left out. So, it seems like the training is reserved for the ‘chosen few’…the unit leaders pick those who are close to them and at times you hear of cases of bribery…”*
>
> — (Male CHV - Kibra)

#### CHV supervision and support

As earlier described, in the Kenyan context, CHVs are supervised by government employed community health assistants (CHAs). The key challenges reported by the CHAs as negatively impacting their ability to offer supportive supervision to CHVs included lack of resources such as transport to facilitate movement within the community especially where there were large areas to cover; and phone airtime for communication. The CHAs were also significantly understaffed which led to increased workloads. Furthermore, one sub-county official explained that recently recruited CHAs had not been adequately trained on the roles of the frontline CHVs, and as such could not offer adequate supervision to the CHVs. Importantly, as illustrated below, the CHAs found it difficult to hold CHVs accountable for expected work, since the role was voluntary, and one could therefore not require them to undertake work.

> *“…the challenge is that CHVs are not given a stipend by the county government, and you see the [implementation] partners cannot give all the CHVs stipends. So, you find some [who are involved in implementation projects], receive stipends while others do not. So, for the ones who are getting, they will really work … they will give you reports. And it becomes easy for accountability purposes, to tag them, onto that stipend… But for those ones who are not getting any stipends, it’s very challenging because for some months they will bring their reports, while for other months they will not. And since you’re not giving them anything, you cannot hold them accountable and ask them for reports. So, I think that is the biggest challenge…”*
>
> — (Sub-county official)

Some of the younger CHAs also found it difficult to supervise CHVs who were older than them, sometimes of the same age as their parents. This was also context-dependent given the prevailing societal norms of showing respect and reverence to one’s elders. Nonetheless, one respondent, a sub-county official, held a differing opinion. According to them, the relationship between young CHAs and the more senior CHVs was not dependent on age, but rather the nature of relationship and interaction between the two groups. That is whenever, the younger CHAs treated the senior CHVs with respect then the respect will be reciprocated back.

#### Supplies and resources

All the participants reported infrequent provision of resources and supplies as a key factor affecting CHV performance. Areas that had ongoing projects implemented by non-governmental partners regularly received necessary resources, whereas other areas did not, which also highlights an issue around (in)equity in resource distribution based on locality and presence of well-resourced partners. The most commonly mentioned resources and supplies that the CHVs lacked and which impacted on their ability to adequately undertake their roles included; reporting booklets, identification documents (badges and/or branded t-shirts), gloves, masks and sanitizers (especially in the context of the COVID-19 pandemic), gumboots and raincoats for the rainy season, and medical cover.

The lack of reporting booklets meant that CHVs often used their own money to photocopy required documents to enable reporting. An important finding given the general low socio-economic status of the study communities of which the CHVs were part of. On the other hand, lack of identification materials made working in some circumstances especially difficult. For example, at the height of the COVID-19 pandemic, the Government of Kenya instituted a dusk to dawn curfew amongst other restrictions. This meant that CHVs could not offer their services during curfew hours, even in the event of an emergency, as they risk being jailed or fined if found outside. Similarly, it made it challenging for CHVs to be prioritized to receive the first round of the COVID-19 vaccine; as they were not considered to be essential workers. The lack of identification also made first-time entry into new households difficult, especially in the context of heightened insecurity in the study communities, sometimes exposing the CHVs to risk of violence from household members. In particular, household members were sometimes suspicious of new, young, male CHVs who were perceived to be spies for thieves in the neighborhood. The quote below illustrates this concern.

> *“…Those who primarily undergo these challenges are the youthful male CHVs. They are not allowed to go into peoples’ houses because of the belief that they are there to spy for the various gangs in the neighbourhood, and then come back later in the night to steal.”*
>
> — (Male CHC representative - Kibera)

Lack of personal protective equipment such as gloves, masks and sanitizers was reported by many respondents as exposing CHVs to risk of infection when conducting household visits, especially during the pandemic period. The CHVs also stated feeling demotivated to continue working during this period due to fear of falling sick, exacerbated by a lack of medical cover and thus not being able to meet hospital costs in case of hospitalization.

> *“…one cannot go and sensitize households about proper ways to wear a mask, practice handwashing and things like that; and in case you fall sick there is no one to turn to…it’s you to sort yourself out…and you know how expensive it is to treat COVID-19 especially when admitted in the ICU.”*
>
> — (Female CHV, Mathare)

#### Remuneration of CHVs (Financial and non-financial incentives)

Given the voluntary nature of the job, CHVs were not always present to carry out their duties as assigned by the CHAs. Since they also needed paid jobs, to provide for their families as outlined below by a CHA.

> *“…Whenever I plan to do an activity with the CHVs in the community, I normally call like 10 CHVs, and book appointments with them. But at the end of the day, I will only get maybe two or three CHVs…Others will ask whether you will compensate them for their time since they have to go out and look for paid jobs…”*
>
> — (CHA, Mathare)

CHVs depended on stipend from NGO trainings & activities to provide for their families. However, the long turn-around payment time demotivated them. According to the county officials CHVs who were called for training/activities with stipend were more active than those called for trainings/activities with no stipend.

Despite the work-related challenges that the CHVs faced, they drew motivation to continue with the work from the appreciation, respect, and recognition that they got from the community. Community members referred to some of the CHVs as doctors, teachers, and good Samaritans. As a result of this respect and recognition, some of the community members visited the CHVs for solutions to the different challenges that they experienced, as shown in the quote below.

> *“…What makes me proud to be a CHV is the respect and recognition I get from the community…you find that when something is wrong in the community, let’s say a child is sick or has refused to go to school, or there are couples fighting, you will be the first person to be consulted by the women in the community…”*
>
> — (Female CHV, Mathare)

### Health system factors

One of the key roles of CHVs in the Kenyan context is to act as a link between the community and the formal health system. A sub-optimal health system and challenges experienced in health facilities by community members, led to loss of community trust in CHVs and made it difficult for them to refer community members to local public health facilities. Some of the key health systems related challenges included poor interactions between healthcare workers and community members at the local referral facilities, unavailability of services, perceived corruption within health facilities, and issues related to referral outside of the CHVs’ local health facilities. All of these resulted in an erosion of community trust in the CHVs as described in detail below.

Most Community Health Committee representatives and CHVs stated that some healthcare workers in public health facilities were rude to patients, resulting in a preference for local private clinics and dispensaries. These sub-optimal interactions between patients and healthcare workers made it difficult for CHVs to refer community members to local public health facilities where they had appropriate linkages as illustrated in the quote below.

> *“…We [CHVs] are supposed to link our clients to government facilities but you find that those in the community prefer going to private facilities, since those [healthcare workers] in government facilities are rude. At times [the patients] go to a public facility with referral letters from a CHV, but the kind of response they will receive…will make them not to go back there…”*
>
> — (Male CHV, Kibera)

Service unavailability was another health system challenge that impacted on CHVs’ ability to perform their roles. For example, most of the government health facilities that were supposed to operate 24-hours a day, did not have operational laboratory and pharmacy services at night. This meant that community members who were referred to these facilities after-hours, had to return the following day for the additional services, causing inconvenience. This in turn led to a loss of community trust in the CHVs who had made the referral in the first place, as seen in the quote below.

> *“The [public] hospitals now work for 24 hours. However, when you refer a patient at night, they will see the doctor but be told to come back the following day for tests and medicine since both the laboratory and the pharmacy are closed for the day. So, the referred patients feel they wasted their time coming to the hospital at night, since they will need to come back the following day for the same service. The community members also think we are in business with the hospital and that we are paid for the referrals…whereas all we are doing is voluntary…”*
>
> — (Female CHV, Mathare)

Another factor that impeded CHVs from optimally undertaking their roles was perceived and actual corruption within the health system. Many CHVs alleged that referred patients paid for goods and services that were supposed to be free at the local public health facilities. These included the child wellness (immunization) booklet and maternity services. The payment for free goods and services in turn led to loss of community trust in the CHVs since the referred patients believed that some of the CHVs were working with corrupt healthcare workers to steal from them. The CHVs also stated that - rather than treat patients at the public facilities which were either free or highly subsidized -some health facility staff allegedly referred community members to private facilities and pharmacies which were owned by the health workers themselves. Thus, the healthcare worker benefitted financially, while the patient incurred an unnecessary out of pocket expense. The illustrative quote below highlights the sale of goods and services that are ideally supposed to be free, and how this in turn impacts on community trust in CHVs.

> *“If it wasn’t for us, these facilities would not have patients…we normally go out campaigning…asking patients to come for free services in government facilities then when they come, they are asked to pay for goods & services that are supposed to be free. For example, expectant mothers are sold for child welfare [immunization] booklets…which cost 200 shillings each… We don’t understand why the booklets are sold while on the back cover it’s indicated ‘Not for sale, Government of Kenya’…this makes the community think that we are working with the facility [health workers] to steal from them*
>
> — (Female CHV, Mathare)

Many of the CHVs were unclear about where their role ended when it came to referrals. For example, it was not clear whether they were required to escort patients to the facility upon referral. Sometimes they escorted patients to facilities outside of their locality where they were unknown by the healthcare workers and ended up experiencing extensive waiting times due to long queues. This in turn demotivated them, and especially given the voluntary nature of the role. Additionally, their referral letters were often not recognized in these facilities outside of their locality; and they were sometimes made to pay for the treatment of the escorted patient as they were perceived to be relatives. All these in turn adversely impacted on CHVs’ ability and willingness to undertake their role of referring patients as illustrated below.

> *“If you accompany a patient to a hospital like Kenyatta [national referral hospital] or send a patient with a referral letter, the healthcare workers there do not recognize the referral letter or even you as the CHV…You will queue with the patient the whole day…they will not recognize that you are a CHV…[it is demotivating]”*
>
> — (Female CHV, Kibera)

### Broader contextual factors

The major broader contextual factors reported to negatively impact on CHVs’ performance and motivation were; demand for financial or material support from community members, perceived corruption in community programmes; and insecurity.

Despite the CHV role being voluntary, some community members perceived CHVs to be government-salaried employees, who benefited from collecting their (household) data. This perception led to CHVs receiving financial and material requests from community members when they visited needy households for data collection. As illustrated below, if they failed to grant these requests, the concern households became uncooperative, which in turn made the work very difficult and demotivating for CHVs.

> *“…at times you may visit a household and find the inhabitants helpless [very impoverished] … You’ll find they’ve had nothing to eat. So, if you visit them today and give them fifty shillings, they will expect you to do the same the next time you visit. And this problem is not only with one household, you might find that there are even 10 households. So, when you visit and tell them that today you have nothing to give, they become uncooperative and start to insult you… At times, you would like to go visit the households but get discouraged since you are there as a volunteer, but the community thinks that you are being paid for the work you are doing.”*
>
> — (Female CHV, Mathare)

Perceived corruption in community programs also adversely impacted CHVs’ motivation and ability to undertake their roles. For example, at the onset of the COVID-19 pandemic, both government and NGOs listed households within the UIS for social support programmes such as cash transfers. However, according to most respondents, the intended beneficiaries never received the payments or material benefits. Since the CHVs had undertaken the household registration exercise, it was assumed that they had benefitted from the programmes at the expense of the community members. This resulted in reputational damage for the CHVs and a loss of community trust in them as illustrated below.

> *“…We faced many challenges during the COVID-19 period… You [CHV] are told to write down names [households], which you do and submit [to the relevant authorities]. But the community members do not benefit thereafter. This has made the community unresponsive and if you now visit households for reports, they refuse to cooperate & tell you that, “you have used our names a lot & it’s you who is benefiting”… This has ruined our name…”*
>
> — (Female CHV, Mathare).

Urban informal settlements are generally characterized by high levels of insecurity that can potentially compromise personal safety. This was similarly the case for the study communities, with respondents stating that the situation was exacerbated by widespread drug use and high levels of unemployment in these contexts. Given this high-risk context that the CHVs worked in, they were constantly at risk of being robbed and having their valuables stolen while undertaking their official roles such as household visits. The female CHVs in particular, were also at risk of being raped and sexually assaulted, especially when working with youths who were drug users; presenting a specific gendered challenge. This emergent theme of the ‘working environment’ and especially concerns about personal safety is a unique challenge that is experienced by CHVs working in urban and peri-urban informal settlements and that has not been noted in the rural and nomadic contexts of Kenya.

> *“…The are a lot of challenges [especially at night] …I am a victim of that…there is a day I came across some thieves [as I was taking a patient to the health facility]. They took pity on me because I had a patient. But they robbed me of everything I had. For the patient, they only touched her inappropriately since she was too sick…if it wasn’t for that, they could have raped her…”*
>
> — (Male CHV, Mathare).

## Discussion

This study explored factors that affect CHV performance in urban informal settlements within Nairobi Kenya and ways in which CHVs can be better supported to enhance their wellbeing and strengthen community strategies. The key programme design factors identified as influencing the performance of CHVs working in UIS included: CHV recruitment; training; the availability of supplies and resources; and the remuneration of CHVs. Health system factors that influenced the CHVs performance included: the nature of relationship between healthcare workers at local referral facilities and community members; availability of services and perceived corruption at the referral facilities; and CHV referral outside of the local health facility. Broader contextual factors that affected CHV performance at the community level included: demand for material or financial support; perceived corruption in community programmes and neighbourhood insecurity. These are discussed in turn below with some suggested recommendations for tackling the issues.

The voluntary nature of the CHV position influenced the kind of CHVs who were recruited and their availability to carry out their duties as assigned by their supervisors. This mirrors findings from other contexts. For example, Sarma et al, George et al, Aseyo et al, and Osindo et al in their work conducted in Bangladesh, India, and Kenya respectively. In Sarma’s work although conducted in a rural setting, the availability of paid assignments made it difficult for the BRAC’s local office to find volunteer workers (47). Whereas, in George, Aseyo and Osindo’s work, CHVs working in UIS found it difficult to devote time to voluntary work without any government allowances. This is because they faced additional pressure to provide for their families and therefore had to look for alternative sources of income. As such, some of them fitted their CHV-related tasks around normal daily income-generating activities (35,48,49).

In Bangladesh, Alam and others noted that a regular salary or income was critical to improving the CHVs’ level of activity (50). Relatedly, to control for attrition, Bhutta et al., recommend regular, performance-based financing (PBF) incentives or hiring CHVs as full-time waged employees (51). Nonetheless, given the significant resource and system challenges experienced in settings such as Kenya, sustainability of such financial incentives are likely to face challenges, including irregular or insufficient disbursement of payments (1). Moreover, evidence from literature suggests that approaches like PBF incentives sometimes result in over reporting (52) or neglect of unpaid tasks by CHVs (2). Therefore, for such incentives to work, there is need for a mix of both financial and non-financial incentives, robust accountability measures, and a consideration of context. It is, however, noteworthy that Nairobi County, Kenya is in the process of establishing a performance-based incentive (PBI) for CHVs. Under this initiative, CHVs will be issued with a monthly stipend of 3,500 Kenya shillings of which 500 shillings will go towards their medical cover. These payments will be in exchange for eight days of work in a month and subject to the CHVs successfully achieving an 80 percent performance target (53). It remains to be seen how this approach will work. Further work is therefore needed to explore the effect of this PBI on the CHVs’ motivation, job satisfaction, attrition, and performance.

Although money was noted as a key motivator for CHVs working in UIS in this study, other non-financial incentives were also observed to play an important role. In particular, CHV appreciation, respect and recognition were seen to be powerful motivators in continuity of the volunteer CHV role. This finding concurs with a review by Jaskiewicz et al., (54) and studies by George et al., Laston et al., and Alam et al (49,50,55,56). In their studies conducted in Ethiopia, Malawi, Zanzibar, Mozambique, Zambia, Nigeria, Burkina Faso, Iran, Pakistan, Nepal (54), India (49) and Bangladesh (50,55,56) these authors found that social prestige and positive community feedback to CHVs were important non-financial factors associated with CHV activity; and should therefore be considered by programmes working in UIS.

As with the study conducted by Kok and colleagues (2), this study found that inconsistent CHV training; inadequate supportive supervision; and lack of, or insufficient, CHV resources and supplies negatively impacted CHV motivation and job satisfaction. This in turn adversely affected CHVs’ wellbeing and performance. This finding concurs with that of several other authors that have conducted work in LMICs including Aseyo et al, George et al, Goudet et al, Kok and Muula, Karuga et al, Laston et al, Martinez et al, Osindo et al, Odhiambo et al, Sarma et al; and systematic reviews by Jaskiewicz et al and Kok et al (2,35,47–49,54,56–61). For example, Aseyo et al., George et al., and Sarma et al., found that inconsistent training left CHVs with knowledge gaps related to the technical aspects of their roles, thus compromising their credibility. This was especially evident when CHVs were not able to respond to (health and other) questions asked by community members (35,47,49).

As in this study, work conducted in other resource-poor settings also show that inadequate supportive supervision of CHVs and insufficient resources and supplies; have also been linked to loss of credibility and subsequent demotivation of CHVs resulting in poor performance (2,35,47,48,54,57,61,62). Previous work in Kenya by Karuga et al., concurred with the findings of this study on the reasons for inadequate supportive supervision (61). Karuga et al found that the main reasons for insufficient supportive supervision were heavy workload on the part of supervisors, inadequate training by trainers who also act as supervisors, and inadequate resource inputs. According to Karuga et al., CHEWs supervised many CHVs, which affected their ability to provide adequate regular supervision. Therefore, to boost CHV supervision in UIS, CHV programmes need to adopt an approach that takes into consideration the trainers’ workload, the voluntary nature of the CHV position, and the limited resources in terms of operational support.

In this study, sub-optimal health systems and related challenges also contributed to loss of credibility for CHVs. This finding resonates with observations by George et al, Sarma et al and a systematic review by Jaskiewicz et al (47,49,54). According to Jaskiewicz and others for instance, community trust or respect was dependent on many factors within the community, with the health facility as an institution having a higher degree of influence (54). Both George et al., and Sarma and colleagues noted that community trust or respect was an important factor to the success of CHV programmes; and when compromised, it rendered CHVs ineffective. As in this study, George, Sarma and colleagues, and the systematic review by Jaskiewicz and others found that sub-optimal healthcare worker-community members interactions, drug stockouts, service unavailability, and perceived corruption in referral facilities; led to loss of community trust in the CHVs. This in turn resulted in a preference by community members for private clinics over government facilities; and subsequently demotivated CHVs since they were evaluated based on referrals made to public health (link) facilities (63,64).

Another health system challenge reported to demotivate and negatively affect CHV work was poor interactions between the CHVs themselves, and referral facility healthcare workers. In this study, this was particularly prevalent when CHVs referred or accompanied community members outside of their usual locality due to lack of clarity on the CHV roles. According to Kok et al., attitude of healthcare workers towards CHVs had a major effect on how they felt and performed (2). Negative attitudes from formal healthcare workers towards CHVs resulted in communication challenges, and adversely affected the community-health facility referral process (65). This suggests a need for awareness raising amongst relevant stakeholders to appreciate the role played by all cadres within the health system, clarity of roles and remit for CHVs, and potentially also communication training for formal healthcare workers to improve interactions with both community members and CHVs.

In this study, the demand for financial or material support, perceived corruption in community programmes, and insecurity hampered CHVs’ service delivery to those living in UIS. These mirror findings from earlier studies conducted in Nairobi and other Kenyan counties by Aseyo et al., Osindo et al., and Odhiambo and colleagues (35,48,58). In these studies, the community expected tangible support from the CHVs. As such, out of sympathy, some of the CHVs used their own financial resources to provide this support to community members, which also led to community acceptance of them. Those CHVs who were unable to meet such demands lost favour especially with needy households, resulting in CHV dropout. This emphasizes the need for clarity of roles including community sensitization of the same (2). This also suggests a need to integrate CHV activities with government social support programmes, that provide assistance to the very needy and shield CHVs from unrealistic and unfeasible community demands. Such social support programs would, however, require proper monitoring and accountability checks to ensure that only genuinely needy community members benefit.

Safety of CHVs especially at night was also noted to be of concern in this study. Indeed, this particular finding seems to be of greater significance in urban informal settlements (including in Kenya) compared to rural and peri-urban contexts. For example, reviews by Glenton et al., and Ogutu et al., (22,66) showed that safety among CHVs working in UIS was of great concern especially due to high poverty levels in these settings, and other social challenges including criminal activities in the local areas. In Brazil for example, CHVs lived and worked in dangerous neighbourhoods with high criminal activities such as prostitution, drug use and violence. Because of this, they felt insecure working in such neighbourhoods for fear of being victims of aggression (67). In previous Kenyan studies it was noted that insecurity hampered the CHVs’ access to certain neighbourhoods or working late in the evenings consequently blocking CHVs from reaching their full potential (48,58). It is therefore pertinent to boost safety and security measures in settings such as where this study took place. These initiatives can include: providing robust and corrupt-free national and local community policing including responsive emergency hotlines as needed; provision of adequate ambulatory services to avoid CHVs having to accompany community members to facilities at night while on foot or motorbikes; tackling social challenges such as high unemployment and drug and alcohol abuse that contribute to crime; ensuring proper and adequate amenities such as local street lighting and good road networks are provided by the relevant national and sub-national authorities; and maintenance of relevant infrastructure. These efforts would require intersectoral and intergovernmental cooperation with both the national and county governments playing a role supported by relevant private and NGO stakeholders.

### Limitations of the study

One key limitation of this study was that it was conducted during a transition phase, when a new CHS (2020–2025) was being rolled out. At the time of conceptualizing the study, this transition had not been anticipated. This meant that the research team had to take into consideration changes in both the new and outdated policy. To mitigate against this, the research team took this change into consideration when designing the data collection tools and factored in interview questions that referred to both the new and outdated policies during discussions.

Additionally, this study was conducted during the COVID-19 pandemic, and subsequently required adherence to COVID-19 safety protocols and restrictions. These always included social distancing and wearing facemasks which was challenging given the study contexts i.e. urban informal settlements. These COVID-19 safety protocols potentially also affected the quality of audio-recordings, especially where the background environments were noisy (given that respondents had to be masked and sit far apart from each other). To mitigate against this challenge, the research team did two interviews for each category of participants. The research team also asked the participants to raise their voices while speaking, and where necessary - especially for soft-spoken participants – to lower their masks only while speaking for audibility. For the soft-spoken participants the interview moderator also repeated their responses to enable capturing by the audio-recorder and to facilitate discussion and responses from others present. The research team also did a thorough quality check of the transcripts as explained in the data analysis section above.

## Conclusion

Like in other settings such as the rural and peri-urban settings, community health volunteers working in UIS in Kenya face a myriad of challenges that impact on their wellbeing and work performance. These cadre of health workers can be better supported by being financially remunerated in addition to other non-financial incentives. Other programme design requirements that are vital for CHVs’ wellbeing include timely holistic training, adequate supportive supervision and sufficient resources and supplies to undertake their job roles. Additionally, there is need for clarity of roles and scope of work, training of healthcare workers on appropriate relations with both the community and CHVs, ensuring availability of services, and putting strategies in place to safeguard against corrupt practices in public health facilities and community programs. In addition, specifically for the urban poor context, there is an urgent need for improved and adequate security measures at the community level, to ensure safety of the CHVs as they undertake their roles.

## Data Availability

Data will be made available upon reasonable request.

## Abbreviations

CHS: Community Health Strategy
CHV: Community health volunteer
CHC: Community health committee
CHU: Community health unit
CHA: Community health assistant
CHEW: Community health extension worker
LMIC: Low-and middle-income country
NGO: Non-governmental organization
FGD: focus group discussion
KII: Key informant discussion

## Acknowledgements

We acknowledge immense support from the Ministry of Health and in particular the Division of Community Health Services. Special thanks to the Nairobi County CHS coordinator and both Mathare/Ruaraka and Lang’ata/Kibra sub-county CHS coordinators for facilitating our community entry and mobilization of both sub-county and community respondents. We also acknowledge the continual support of the Nairobi County Department of Health, and all the study participants for giving their time during the study period.

## Authors Contributions

Michael Ogutu (MO), Timothy Abuya (TA) and Kui Muraya (KM) conceptualized and designed the study. MO with support from KM developed the data collection tools, led the data collection, cleaning and analysis; and drafted the initial manuscript. Erick Kamui (EK) assisted with the data collection and preliminary analysis. KM supported and gave overall guidance on both the data collection and analysis. All authors reviewed the manuscript and gave critical feedback that contributed to its revision. All the authors read and approved the final manuscript.

## Declarations

### Consent for publication

This paper is published with the permission of the Director, Kenya Medical Research Institute (KEMRI).

